# Social relationships and activities following elimination of SARS-CoV-2: a qualitative cross-sectional study

**DOI:** 10.1101/2021.09.20.21263837

**Authors:** Nicholas J. Long, Nayantara Sheoran Appleton, Sharyn Graham Davies, Antje Deckert, Edmond Fehoko, Eleanor Holroyd, Nelly Martin-Anatias, Rogena Sterling, Susanna Trnka, Laumua Tunufa’i

**Affiliations:** Department of Anthropology, London School of Economics and Political Science, London, United Kingdom; Centre for Science in Society, Victoria University of Wellington, Aotearoa New Zealand; School of Languages, Literatures, Cultures, and Linguistics, Monash University, Melbourne, Australia, and School of Social Sciences and Public Policy, Auckland University of Technology, Auckland, Aotearoa New Zealand; School of Social Sciences and Public Policy, Auckland University of Technology, Auckland, Aotearoa New Zealand; School of Māori Studies and Pacific Studies, University of Auckland, Auckland, Aotearoa New Zealand; School of Clinical Sciences, Auckland University of Technology, Auckland, Aotearoa New Zealand; University of Waikato, Hamilton, Aotearoa New Zealand; Department of Anthropology, University of Auckland, Auckland, Aotearoa New Zealand

**Keywords:** COVID-19, elimination, health policy, pandemic response, New Zealand, social isolation, social recovery

## Abstract

**Objectives:** To investigate how successfully SARS-CoV-2 elimination strategies fulfil their promise of allowing a return to a ‘normal’ social life, and to identify obstacles and challenges that may inhibit the realisation of this goal.

**Design:** Qualitative cross-sectional survey.

**Setting:** New Zealand community cohort.

**Participants:** 1040 respondents entered the study (18–90 years, M = 48.18.11, SD = 15.52, 76% women). 966 completed the questions relevant to this article. Participants were recruited via online advertisement campaigns designed to maximise variation in the sample as far as practicably possible.

**Main outcome measures:** Thematic analysis of participants’ narratives.

**Results:** A majority of participants reported that the elimination of SARS-CoV-2 had allowed their life to go back to being ‘more or less the same’ as before the pandemic. A small number indicated the pandemic had inspired them to become more social following elimination. Nevertheless, a sizeable minority of respondents reported being less social, even many months after SARS-CoV-2 had been eliminated. This was often because of fears that the virus might be circulating undetected, or because the March-May 2020 lockdown had led to changes in relationships and personal habits that were not easily reversed. Becoming less social was associated with having an underlying health condition that heightened one’s vulnerability to COVID-19 (*p* = 0.00005) and older age (*p* = 0.007).

**Conclusions:** Elimination strategies can successfully allow the public to return to a pre-pandemic ‘normal’ – or reinvent and improve their social lives should they wish. However, such outcomes are not inevitable. Re-establishing social connections after elimination can sometimes be a challenging process, with which people may need support. Plans for providing such support should be an integral part of elimination strategies.

## Introduction

Since the onset of the COVID-19 pandemic, politicians and public health experts have disagreed over how to respond to outbreaks of SARS-CoV-2. With few prepared to let the virus spread unimpeded, debate centres on the respective merits of mitigation and elimination strategies. In mitigation, the virus continues to circulate at reduced levels, due to non-pharmaceutical interventions (NPIs), which may either be undertaken optatively by the public or legally imposed. Elimination, sometimes referred to as ‘Zero-COVID’ – involves the stringent deployment of NPIs with a view to reducing community spread to zero; further outbreaks are then guarded against via strict border controls, at least until appropriate vaccination thresholds are reached.^1^

Notwithstanding technical questions regarding the feasibility of elimination, which may prove difficult in non-OECD countries or when faced with SARS-CoV-2 strains of heightened transmissibility, proponents of each strategy are motivated by competing understandings of how best to protect the public good. Advocates of mitigation argue that the economic damage wrought by stringent lockdowns and the border closures necessary to secure Zero-COVID status would have long-term repercussions for public health.^2^ Supporters of elimination counter that it minimises COVID-19 fatalities and post-COVID syndrome (or ‘Long COVID’), supports economic recovery by allowing businesses to resume relatively normal service and consumer confidence to return, and ultimately results in less restriction of civil liberties.^3^ Elimination is also seen as having social and psychological benefits, often couched in the idiom of ‘normality’.^4 5^ Not only would endemic circulation of COVID-19, even at suppressed levels, entail ‘profound cultural adjustment for the life of high-risk individuals in the winter months’;^6^ there is, McKee argues in his critique of mitigation, ‘little point in removing restrictions if a large proportion of the population is too worried to place themselves at a real or perceived risk’.^7^ By contrast, elimination is seen as allowing ‘normal social life to resume’^8^ and facilitating ‘largely normal community life’.^9^ Embedded within these claims is a recognition, firstly, that the disruption of established routines is itself a psychological stressor,^10^ and, secondly, that the attenuated social life possible under NPIs can deprive people of much-needed social support, with potentially adverse consequences for physical health, mental health, and subjective wellbeing.^11-13^

To date, empirical evidence supports both the public health and economic arguments in favour of elimination. Zero-COVID strategies are associated with fewer excess deaths than mitigation,^14-16^ while nations that rapidly eliminated the virus have, to date, performed better economically than those in which it was suppressed.^3 17 18^ However, while research shows that people living in settings adopting mitigation strategies are tending to have fewer social contacts than they did pre-pandemic after ‘lockdown’ restrictions have been lifted,^19 20^ little is yet known about the extent to which the promise of ‘normal’ social and community life has been realised following the elimination of SARS-CoV-2. Quantitative analysis of the Gallup World Poll presents a mixed picture, indicating that elimination settings observed an uptick in ‘prosocial activity’ (making a donation, volunteering, or helping a stranger in the past month) but a reduction in reported levels of social support (whether the respondent has someone to count on in times of trouble).^21^ Yet such findings reveal little about the character of everyday social life or the degree to which it feels satisfying, let alone ‘normal’. This study develops our understanding of elimination approaches by qualitatively analysing accounts of life in ‘Zero-COVID’ New Zealand (specifically, between February-August 2021), examining how successfully research participants were able to return to their pre-pandemic ways of life, and identifying specific challenges that governments adopting an elimination strategy should anticipate in future.

## Methods

### Theoretical orientation

Understanding the extent to which elimination allows people to resume a satisfying, ‘normal’, social life requires a qualitative, social constructionist approach. As health researchers have long recognised, a sense of ‘normality’ principally inheres in subjective evaluations of how a practice or situation feels and how it compares to imagined baselines of ‘normal’ activity and behaviour.^22 23^ These evaluations are then expressed through narrative in a process sometimes referred to as ‘narrative sense-making’.^24 25^ Moreover, such evaluations, and their narrative expression, are enacted by individuals whose expectations, understandings, and yardsticks of measurement are both constantly evolving and distinct from those held by others, even as they are to some degree co-constructed between people and within communities and are responses to shared circumstances.^26 27^ Each narrative must thus be engaged with qualitatively, and on its own terms, whilst also remaining attentive to patterns across different cases. These goals can be achieved via the qualitative technique of thematic analysis.^28^

### Site selection

New Zealand is an ideal site in which to investigate social life following the elimination of SARS-CoV-2. The New Zealand government has highlighted ‘get[ting] back to a sense of normality’ as a reason to pursue its elimination strategy,^29^ and the country is frequently assumed to have enjoyed ‘normal’ social and community life following the elimination.^8 9^ Indeed, this is one reason for New Zealand to be widely regarded as a poster child for Zero-COVID approaches.

Elimination first occurred in May 2020, following a 49-day national lockdown,^30 31^ and on 8 June 2020 the whole of the country moved to ‘Alert Level 1’, in which activities were permitted to operate without restriction – although enhanced record-keeping was recommended, and mask-wearing on public transport mandated. Small community outbreaks in August 2020 and February 2021 led to short periods of enhanced restriction, mostly in the city of Auckland. Nevertheless, until the incursion of the Delta variant triggered a further nationwide lockdown in August 2021, New Zealand had enjoyed many months of freedom from COVID-19 restrictions (see Figures 1 and 2). Vaccine rollout began during this time, with frontline workers vaccinated from February 2021 and members of the public vaccinated from May 2021. However, the pace of rollout has been slow, with only 18% of the population fully vaccinated and 29% having received one dose by the start of August 2021.^32^ For most people, the elimination strategy has been the principal line of defence since the pandemic began.

**Figure 1.**
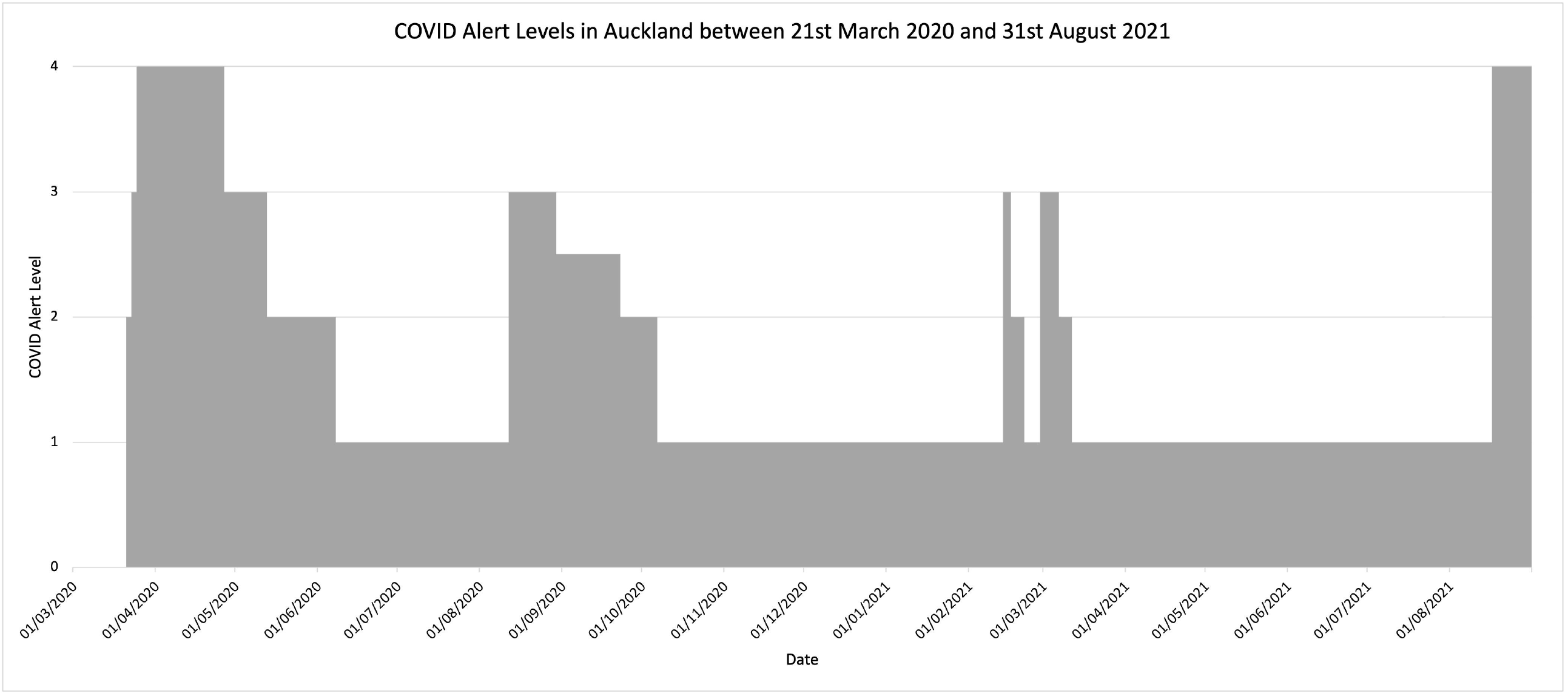
COVID Alert Levels in Auckland between 21st March 2020 and 31st August 2021.

**Figure 2.**
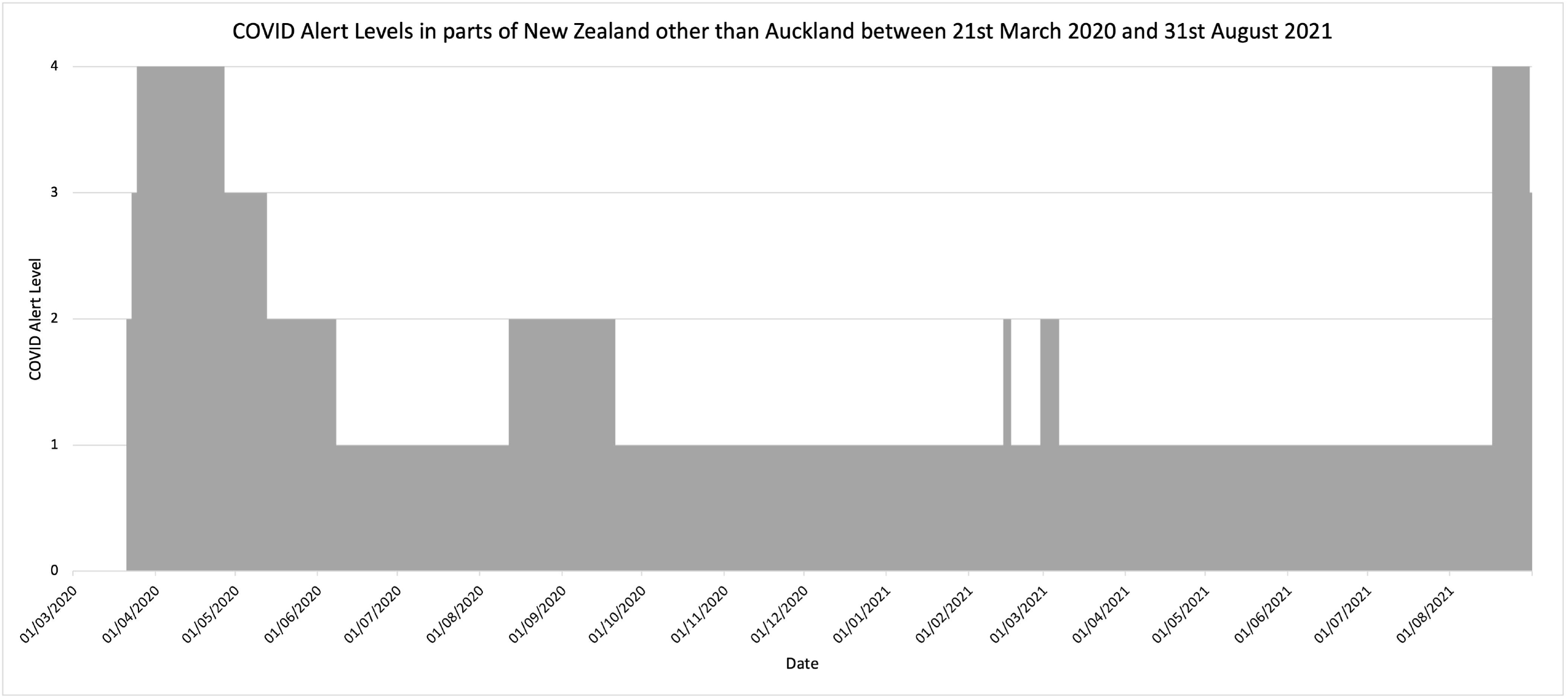
COVID Alert Levels in most parts of New Zealand other than Auckland between 21st March 2020 and 31st August 2021. N.B. Wellington was also at Alert Level 2 for six days between 23 and 29 June 2021.

### Participants

This research was envisaged as a qualitative study. It did not seek to provide a statistically representative account. Rather, following Cole and Knowles’ argument that every ‘exploration of an individual life-in-context brings us that much closer to understanding the complexities of lives in communities’,^33^ it investigated the *range* of possible experiences that people could have following New Zealand’s re-opening, using respondents’ own words to identify key dynamics underpinning different behavioural pathways. Sampling aimed to maximise variation as far as practicably possible, so we could trace contrasts and patterns within the data obtained.^34^ It continued until we observed saturation in the data.

Recognising the flexibility, scalability, and richness of online surveys as a method of gathering qualitative data,^35^ we advertised an online research survey via nationwide Facebook and Instagram campaigns, including bespoke campaigns targeted at men and younger age groups, to heighten variation within the sample. Participants were also recruited from a database of contacts who had participated in the research team’s previous surveys on experiences of the COVID-19 pandemic in New Zealand – who were themselves recruited via advertising campaigns intended to maximise variation.^36 37^ The survey was self-administered and thus unlikely to have been influenced by researcher characteristics. In total, the survey received 1040 valid responses. The respondent pool showed good variation in terms of age and region of residence but, despite our attempts to maximise variation, and as is often the case with survey research in New Zealand,^38^ contained disproportionate numbers of women, New Zealand European / Pākehā people, and university graduates.

### Survey design

We distributed the online survey between 18^th^ August and 25^th^ August 2021, canvassing respondents’ opinions on various aspects of New Zealand’s pandemic response, and assessments of how their lives over the previous six months (including domestic and neighbourhood relations, friendships, social life, working practices and outlook on life) compared to their lives before the pandemic (see Supplementary Annex for a full schedule of questions). Respondents could indicate that these were ‘more or less the same’, ‘a little different’ or ‘extremely different’ and were then prompted to explain their answer in their own words. 966 respondents answered the question about how much their social lives had changed. 494 provided narrative elaborations.

### Analysis

Having anonymised and read these narrative answers multiple times, two researchers (NJL and SGD) independently coded them for analysis under both anticipated themes (e.g., whether they described upturns, downturns or continuity in the quality and quantity of social relationships and activities) and emergent themes discovered through exploring the data (e.g., references to relationships with people overseas, increased use of technology, and epidemiological considerations).^39^ Independent coding helped to ensure trustworthiness. Disagreements over codes were resolved via consensus discussion. We used constant comparison to ensure that our thematic analysis provided a comprehensive overview of themes and subthemes evident in the data.^40 41^

Our coding strategy allowed us to conduct a statistical exploration of patterns in the data.^42^ Using Microsoft Excel v.16.52, we conducted Chi-squared tests for independence to examine the frequency of three key themes – ‘returning to normal’ (code N), ‘becoming more social’ (code P), and ‘becoming less social’ (code L) – across five demographic variables: gender, age, ethnicity, education status, household size, and presence or absence of underlying health conditions that might affect one’s vulnerability to COVID-19. In cases where responses had received multiple codes, we identified a single ‘predominant’ code for the purposes of statistical analysis, since the Chi-square test requires independent observations. Recognising that respondents may have been less likely to elaborate on an answer indicating that their lives were ‘more or less the same’ than when having reported a difference, separate tests were run for associations between the prevalence of P and L codes not only vis-à-vis N codes but also vis-à-vis N codes *and* unelaborated ‘more or less the same’ answers. Since respondents had been able to select multiple ethnic labels, the tests for ethnicity were based on concatenations of the data: we ran separate tests to see whether there were any associations between survey responses and identifying exclusively as White (whether by checking the ‘New Zealand European / Pākehā’ box, or reporting another Caucasian ethnicity under ‘Other’) or identifying non-exclusively as Māori. We also tested to see whether there was any association with being Māori *or* Pacific, since these two groups have been identified as especially vulnerable to COVID-19.^43^ Small cell sizes precluded tests for associations with other ethnicity options in the survey.

### Patient and public involvement

Neither patients nor the public were involved in the design or conduct of this research.

## Results

Thematic analysis revealed three overarching patterns of social behaviour in the wake of elimination. The most common response was that life was ‘more or less the same’ as it had been before the pandemic. However, a minority of our sample reported having become more social, and an even more sizeable minority reported having become less social. These responses varied by gender and age, but also, and most dramatically, by health status, indicating that those who are most vulnerable to COVID-19 may also be least able to achieve social recovery once SARS-CoV-2 has been eliminated.

### Returning to the pre-pandemic ‘normal’

Of the 966 respondents who answered the question about friendships and social life, 531 indicated that these had been ‘more or less the same’ over the previous six months (i.e. from February – August 2021) as they had before the pandemic. When elaborating on their answers, most of these respondents attested that nothing had changed (Table 1 - Quote 1). They described Level 1 as allowing a return to normality (Quote 2), and feeling grateful and ‘lucky’ that the New Zealand government had adopted an elimination strategy (Quotes 3 and 4). Some explained that the social gains of life at Levels 1 and 2 justified the ‘sacrifice’ of lockdowns (Quote 5). Several mentioned that elimination had allowed them to feel ‘safe’ (Quotes 3 and 6) alleviating their feelings of ‘fear’ (Quote 7). One respondent, who had spent three months of 2021 in the United Kingdom – a country which has adopted a mitigation strategy – contrasted the ‘normality’ of Zero-COVID with the ‘frightening’ feeling of socialising post-lockdown in the UK (Quote 8). Only in a handful of cases was the ‘normality’ of social life linked to a wilful blindness towards the pandemic (Quote 9). Interestingly, although most respondents were supportive of the vaccination programme elsewhere in the survey, none mentioned it as a factor contributing to their experience of social recovery.

**Table 1.** Returning to pre-pandemic normality.

|  |  |  |
| --- | --- | --- |
| Quote 1 | My friendships and social life were the same before and after lockdown. My social circle is small anyway so it is not difficult to maintain. | Māori woman, 40s |
| Quote 2 | Things returned to normal once we returned to level 1. | Pākehā woman, 30s |
| Quote 3 | Thanks to the government keeping us safe, we've been able to enjoy most of what we usually do. | Pākehā woman, 60s |
| Quote 4 | We are lucky to have an elimination strategy, so apart from the initial lockdown itself very little had to change. | Pākehā woman, 30s |
| Quote 5 | Every time we are at level 1 or 2 then all of our social life explodes again which is lovely - I'm willing to sacrifice here and there in a level 3 or 4 in order for the huge gains we get the rest of the time. We've been able to take holidays around NZ, go to arts and sports events, I've been able to sing in my symphonic choir with audiences of nearly 3,000, go to big birthday events, rugby, parties, and big work events. It's an incredibly fortunate life so far. | Pākehā woman, 50s |
| Quote 6 | Things appeared to be normal and we felt safe | Pākehā man, 60s |
| Quote 7 | Having the freedom of choice to again visit friends and family without any fear and go have a normal social life. | Pākehā woman, 70s |
| Quote 8 | In NZ everything was normal till this new lockdown so normal life prevailed. In the UK everything was different - unable to see my friends until the UK lockdown relaxed. Even then I was frightened of putting folk in danger. Kept wearing my mask | Pākehā woman, 60s |
| Quote 9 | By turning off all media life can carry on without all the lies and one sided rubbish | Māori and Pākehā man, 50s |
| Quote 10 | I am more mindful of sickness/illness and how my family may impact the health of others and vice versa. I am much more conscious about my movements and that of my family. | Māori and Pākehā woman, 30s |
| Quote 11 | More or less the same but cognizant of public health precautions - washing hands, etc | Pākehā man, 60s |
| Quote 12 | We don't have a raging social life with a young child and a baby on the way. But we've been happy to go to theatres or cinemas. I don't have any friends who are border workers and don't get out too much in level 2 anyway. | European woman, 40s |
| Quote 13 | Although some of us have different opinions about the govt. and their decisions re pandemic, our friendships are strong and we can have robust discussions without getting personal. | Pākehā woman, 60s |

11 respondents, including a few who indicated that their social life and friendships had become ‘a little different’, described minor changes resulting from heightened awareness of health and hygiene, as opposed to substantive shifts in the frequency or character of social activity (Quotes 10 and 11). Nevertheless, several reports of a ‘return to normality’ were haunted by a sense of contingency, with respondents indicating that life might have been less normal if they had friends who were border workers (Quote 12), or if they were less adept at handling differences of opinion within their relationships (Quote 13). This indicates a recognition that the post-elimination context may put pressures on certain relationships.

### Becoming more social

95 respondents shared narratives describing an intensification of social activities following the elimination of COVID-19. Some framed this as a response to the lockdown, couched in the idiom of ‘making up for lost time’ (Table 2 - Quote 1), while other respondents explained that the pandemic had revealed the fragility of social freedoms they had previously taken for granted (Quote 2), and, indeed, human life (Quote 3), inspiring them to prioritise friendships and social activities more than they previously had (Quotes 4 and 5), and to appreciate their loved ones more (Quote 6).

**Table 2.** Becoming more social

|  |  |  |
| --- | --- | --- |
| Quote 1 | I have been going out as much as possible when not in lockdown to make up for lost time | Asian and Pākehā woman, 20s |
| Quote 2 | I think we have embraced our friends and social life with much more gusto, cos you just don't know when we'll be locked down again. We've been doing quite a lot more domestic travelling and local activities. Also trying to support local businesses. | Australian woman, 20s |
| Quote 3 | More social events as don't want to miss out because people could die anytime. | Pākehā woman, 20s |
| Quote 4 | I have made more effort to join in | Pākehā man, 60s |
| Quote 5 | I've built stronger friendships with my flatmates and been far more willing to go out and do things. I think this has partially been in an effort to make the most of the time we are not in lockdown. Prior to the pandemic I often turned down social events to study. | Pākehā woman, 20s |
| Quote 6 | More appreciative of friends who can keep company through hard things like lockdown. Some older friendships refreshed because of time and drive to reconnect - rather than 'leaving it to another day'. | Pākehā woman, 50s |
| Quote 7 | Our neighbourhood formed closer friendships during previous lockdown (socially distanced afternoon teas on the street), and we have continued supportive closer relationships. | Pākehā woman, 60s |
| Quote 8 | Got closer to most people. Am more honest about my emotions | Pākehā woman, 50s |
| Quote 9 | I think my social life has been better with more meaningful relationships | Pākehā woman, 30s |

In several cases, the experience of being ‘locked down’ in 2020 had afforded opportunities for new friendships to be made – especially with those in the local community (Quote 7) – and these new intimacies had persisted into the post-elimination period. Others noted that the challenges of the pandemic had allowed them to forge closer relationships with others, by allowing them to be more honest about their emotions (Quote 8), or by rendering friendships more ‘meaningful’ (Quote 9).

### Becoming less social

253 respondents gave qualitative answers in which life in a Zero-COVID New Zealand was associated with a decline in the quantity or quality of social relationships. In 24 cases, this was explicitly linked to the border closures integral to the elimination strategy (Table 3 - Quote 1). The majority described their social lives and social networks *within New Zealand* having changed since pre-pandemic times. They often reported changes in activities (‘socialising less’ or spending less time in public places – Quotes 2 and 3), changes in character (becoming ‘less social’ – Quote 4), and an overall sense of their world ‘having shrunk’ (Quote 5).

**Table 3.**
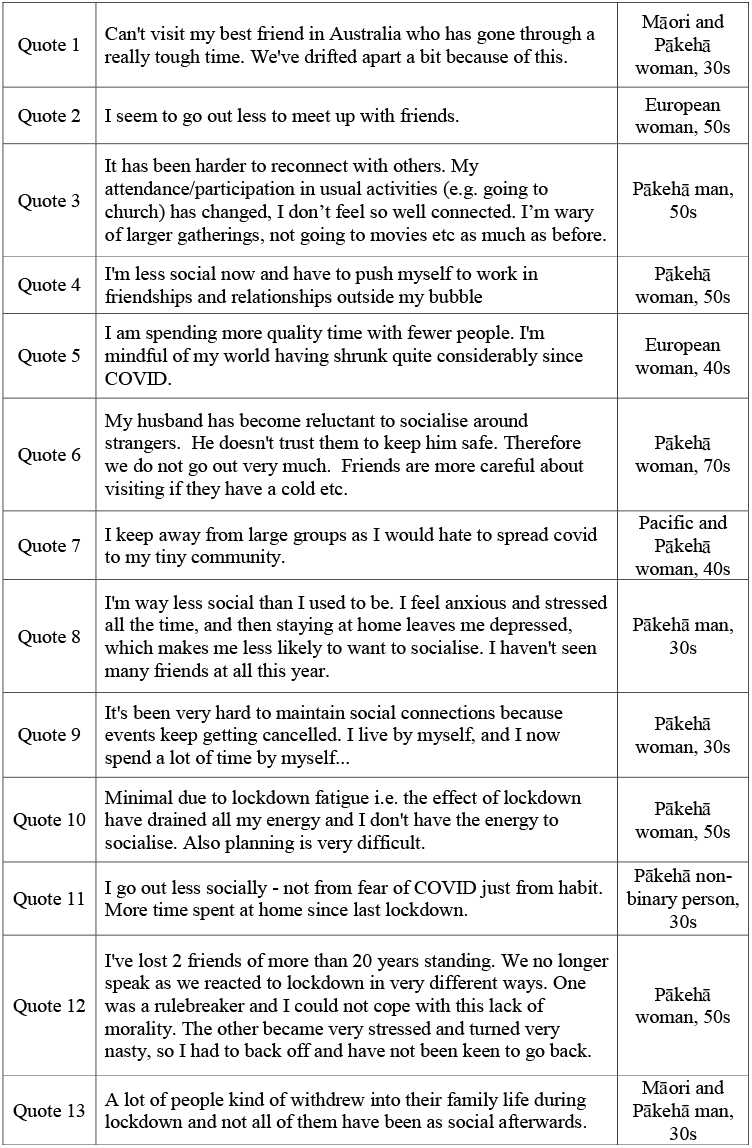
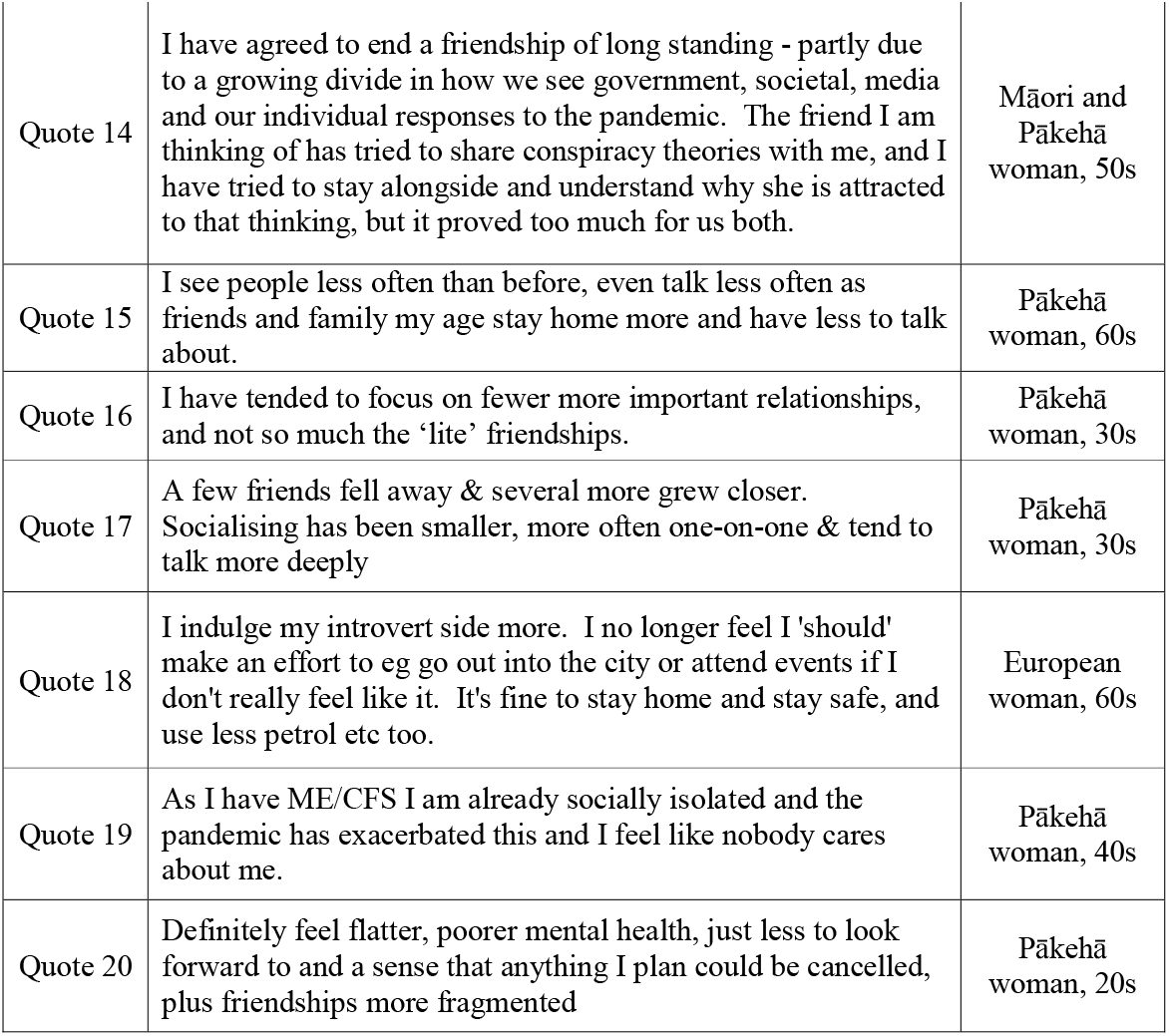
Becoming less social

For some respondents, these changes were linked to an ongoing fear that SARS-CoV-2 might have entered New Zealand and be in circulation, despite announcements that community transmission had been eliminated. They worried that, by socialising, they might either contract it (Quote 6) or pass it to others (Quote 7). Others described how the anxiety and stress that they had experienced due to the pandemic had triggered feelings of depression, that then impeded them from undertaking social activities that they might otherwise want to (Quote 8). Such anxieties could have knock-on consequences, with a diminished social life activity sometimes arising from frequent cancellations (Quote 9).

In other cases, the change in social patterns was presented as a consequence of having lived through the seven-week lockdown in March-May 2020. Some respondents described ongoing ‘lockdown fatigue’ (Quote 10), while others explained how the lockdown had ‘habituated’ them to staying at home (Quote 11). Some friendships had been strained during the lockdown period due to disagreements over the meaning or importance of lockdown rules (Quote 12), while other respondents felt that the lockdown had led people to ‘withdraw into family life’ at the expense of their friendships (Quote 13). Both of these dynamics were also reported as having arisen even after lockdown had ended: disagreements over vaccine uptake or the government’s COVID response had caused some people to sever ties with certain friends (Quote 14), while those whose friends had a propensity to ‘stay at home’ – for whatever reason – noted that those friendships now felt thinner, with less to talk about (Quote 15).

Not all respondents viewed these changes in their social lives as negative. Some appreciated being able to focus only on their ‘most important’ relationships (Quotes 3 and 16), the ‘deep’ conversation afforded by smaller gatherings (Quote 17), or being able to ‘indulge their introvert side’ (Quote 18). For others, however, the loss of connection had led to a heightened sense of isolation (Quote 19) and deteriorating mental health (Quote 20).

### Response Distribution

The three patterns described above could be observed amongst respondents of all backgrounds. Statistical analysis of the prevalence of codes indicating ‘becoming more social’ or ‘becoming less social’ relative to codes or answers indicating a return to pre-pandemic normality (Table 4) did not indicate any significant associations with ethnicity, education status or residence size. There were, however, statistically significant associations with health status and age. Respondents with underlying conditions were more likely to have become less social and less likely to have become more social than those who did not (*p =* 0.00005). Similarly, those in younger age brackets were more likely to be pro-social, and those in older age brackets more likely to be less social (*p* = 0.009) – although this pattern may partly reflect the increased prevalence of underlying health conditions in older age groups. Analysis also revealed a statistically significant association with gender: women were more likely to report having become more social *or* having become less social, and men more likely to report continuity (*p* = 0.007). This finding may reflect gendered differences in behavioural pathways following elimination, but may also reflect longstanding gender roles in Western societies, in which women have often been deemed responsible for thinking about and managing social relationships,^44-46^ and thus potentially more inclined to detect and report changes in their social networks.

**Table 4:**
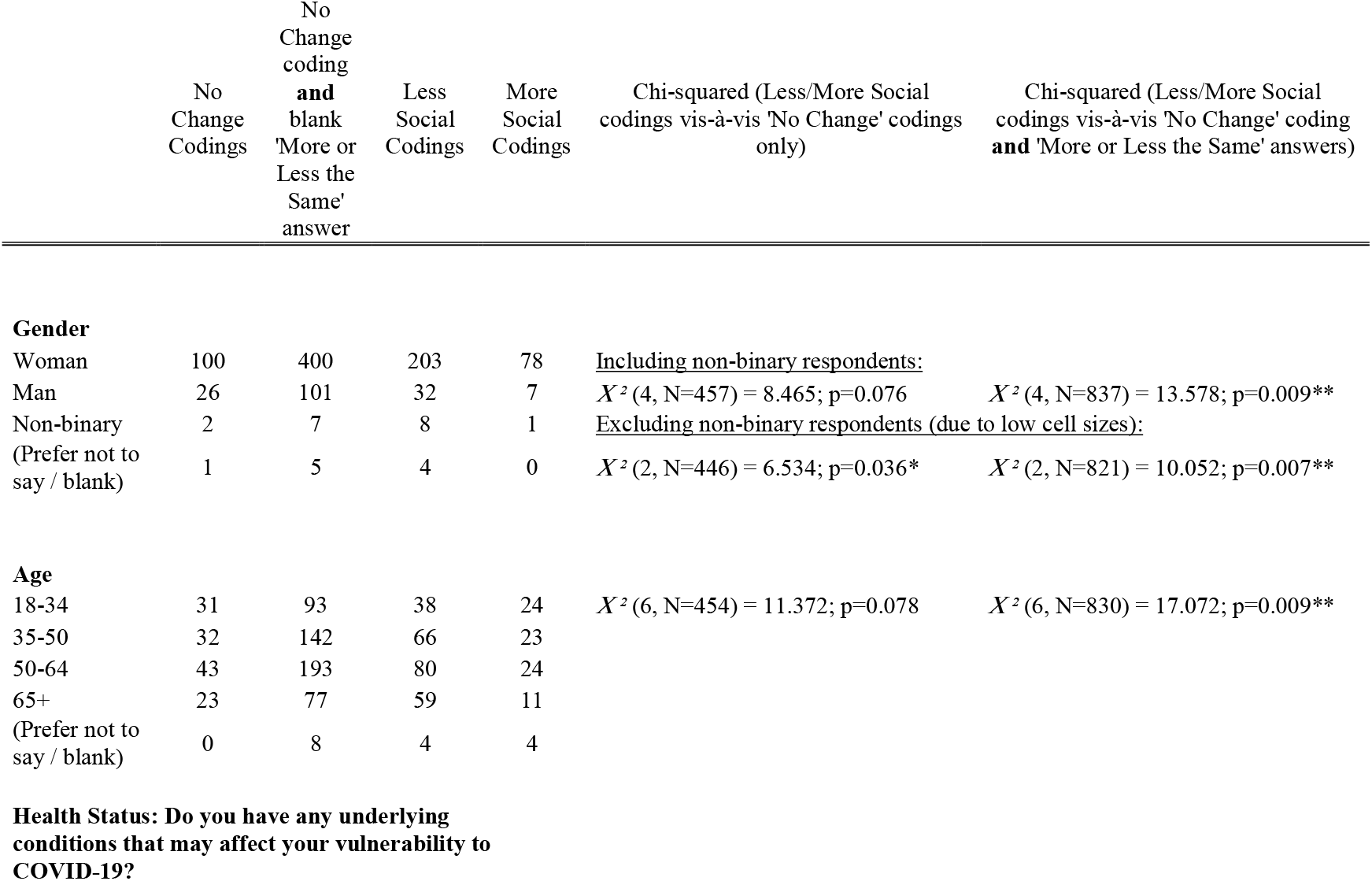
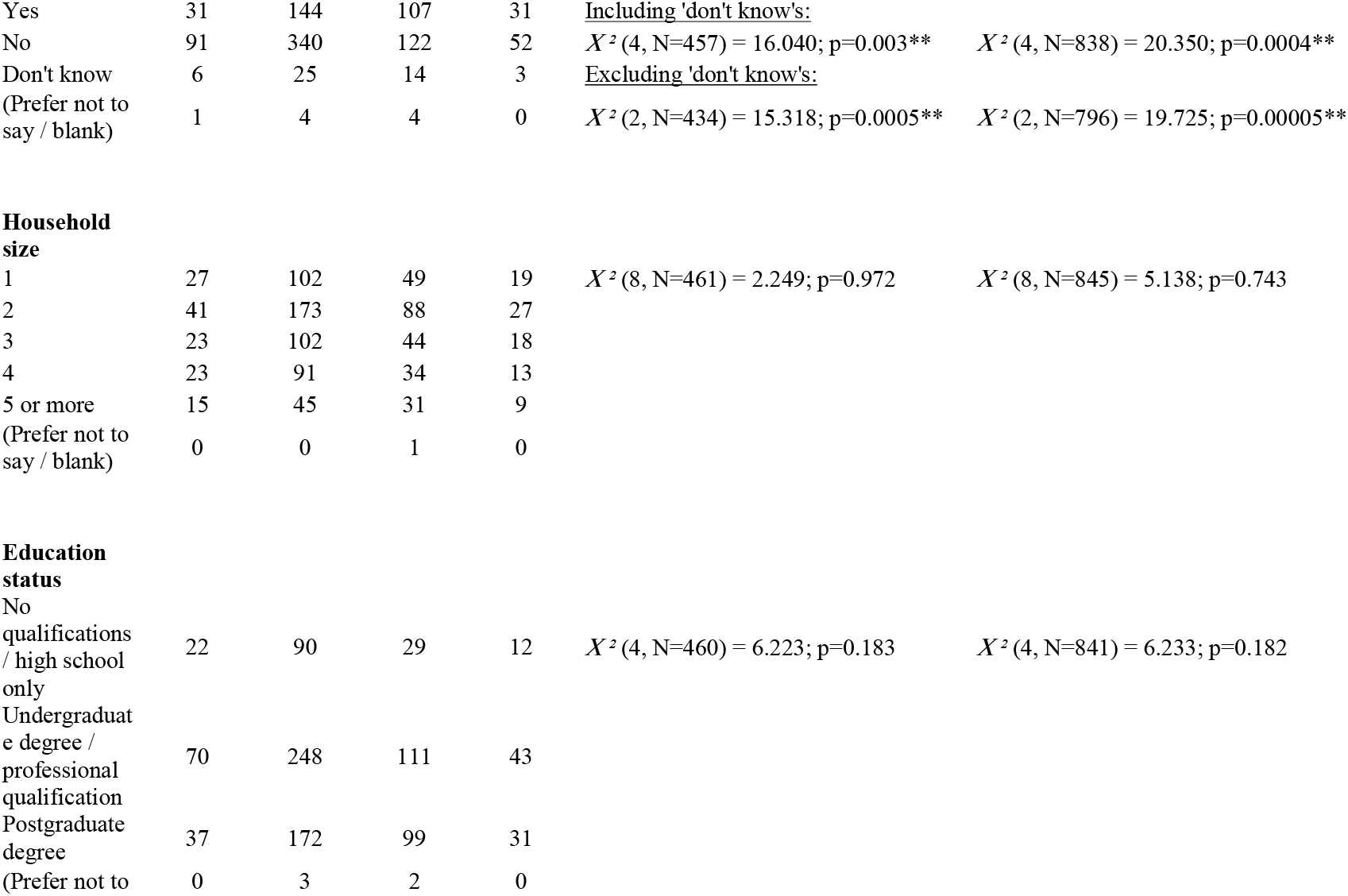
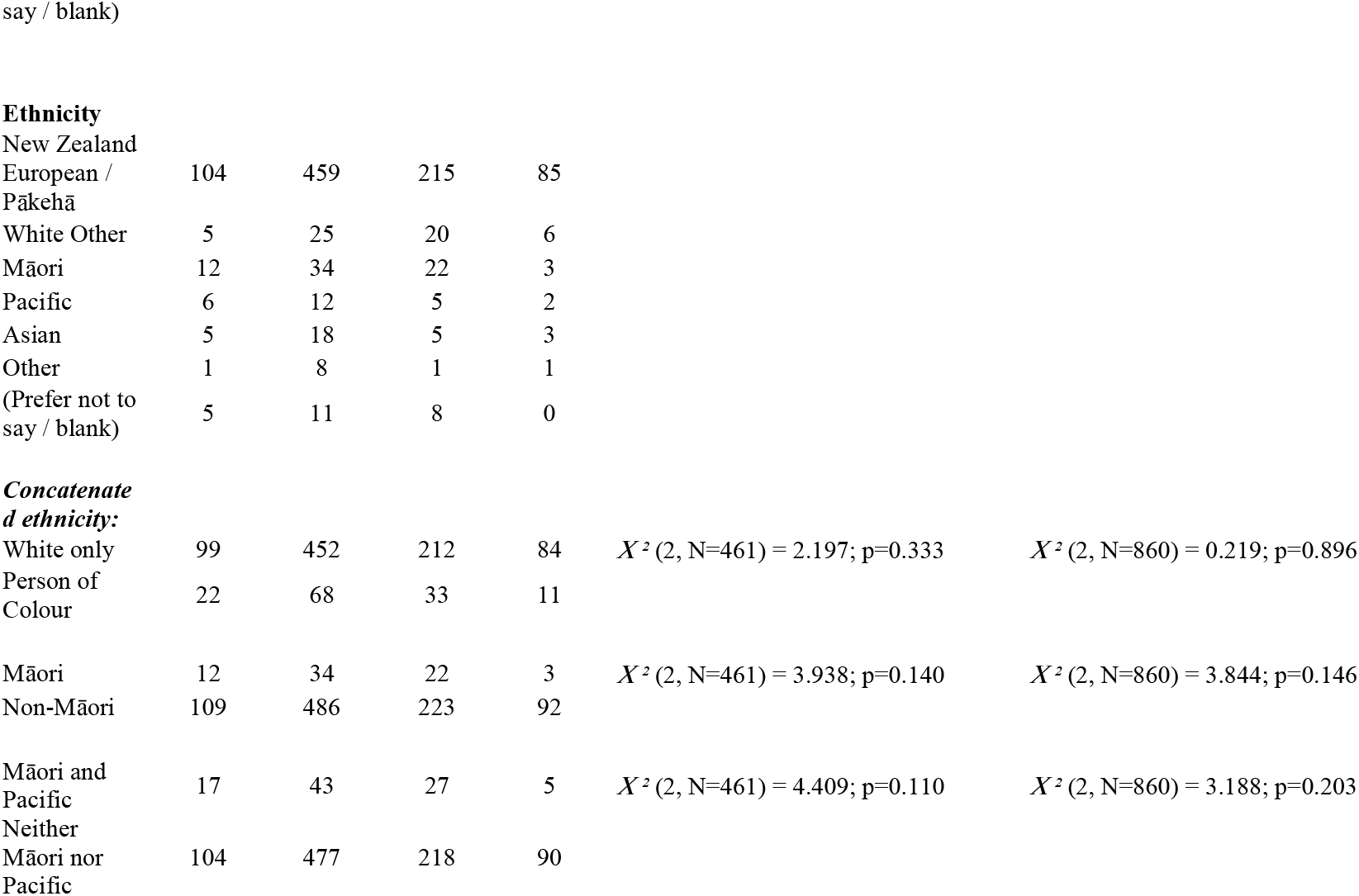
Distribution of responses to question about changes to friendships and social life

## Discussion

Elimination strategies can indeed allow many people to regain a sense of ‘normality’ within their social lives, even inspiring them to adopt modes of sociality they considered better than those they had experienced before. Such positive outcomes, however, are far from inevitable. Many respondents reported lower levels of social contact after the virus had been eliminated than before the pandemic. This was especially, but not only, the case for older respondents and those with underlying health conditions.

While the thematic analysis allowed us to identify three distinct patterns of response to elimination, and key factors that influenced each of those responses, we cannot be certain how well the prevalence of those responses within our sample reflects their prevalence amongst New Zealand’s public. Given the high proportion of respondents identifying as women (76.6%) and reporting underlying health conditions (31.7%), and a mean age above the national average, it seems likely that the pathway of ‘becoming less social’ is overrepresented in our results. Although a statistically representative study would better delineate the scale of the challenges, this qualitative study nevertheless identifies several distinct dynamics that can obstruct ‘returning to normality’, showing a clear need for policy measures and messaging that can support the public in their transition to post-elimination life.

Retrospective self-reporting is sometimes considered inferior to observational studies.^47^ We see the retrospective nature of our survey as a strength, since it allows us to better understand the narrative sense-making undertaken by our respondents. Nevertheless, the cross-sectional nature of the study meant such narrativisation was occurring at a specific moment in time: by happenstance, community outbreaks of the Delta variant led to New Zealand entering Level 4 lockdown just as our survey was due to be launched. The sense of pessimism surrounding the lockdown may have led some respondents to exaggerate what had been lost since before the pandemic, and others to romanticise life immediately prior to lockdown - although a strength of this timing was that it allowed respondents to reflect back on the longest possible sustained period at Level 1. Our study thus complements existing studies on social attitudes and behaviours in the immediate aftermath of the 2020 lockdown, and shows how many of the trends reported in that research, including an aversion to mixing with strangers,^48^ ongoing uncertainty about the trajectory of the pandemic,^49^ and worsening mental health,^50 51^ appear to have persisted for many months, despite minimal domestic COVID-19 cases. It also goes beyond those studies by offering a more integrated understanding of how respondents are experiencing and evaluating their social life, engaging with respondents’ accounts of the pandemic on their own terms, and identifying age and medical vulnerability to COVID-19 as risk factors for social disconnection.

As our thematic analysis reveals, the shrinkage of a social network is not always undesirable: it need not equate to ‘loneliness’, and may even be experienced as a relief. Nevertheless, research in New Zealand^52-55^ and beyond^56-58^ points to strong correlations between the number and quality of social relationships and overall physical and mental health. There are also known psychological benefits associated with living in a world that feels ‘normal’.^10^ Enabling people to restore or expand their pre-pandemic social networks is thus not only a sociocultural prerogative, but also a public health imperative.^59^

Elimination strategies would hence be improved by anticipating and mitigating against common obstacles to people taking full advantage of the opportunities afforded by a Zero-COVID environment. Foremost is the ongoing fear of contagion, which is not necessarily eliminated with the virus – especially when a pandemic continues to rage internationally. In addressing such fear, policy makers must strike a delicate balance between maintaining appropriate levels of caution (e.g., via diligent use of contact tracing, etc.) and encouraging people to take advantage of their hard-won freedoms. Public health messaging should champion adjustments made to make public venues as COVID-secure as possible (for example, by increasing airflow), promote low-risk ways in which one can gather with friends (such as by meeting outdoors), and emphasise that reconnecting with others is a public health good in its own right, perhaps harnessing the tropes of ‘kindness’ and ‘togetherness’ that underpinned New Zealand’s initial messaging around lockdown.^60^

Our study also demonstrates how even a relatively short (49 day) lockdown, especially one of the stringency required to achieve elimination, can have long-term impacts on personal habits, relationships, and mental health – all of which can affect social networks. Funding of mental health services – including both individual psychotherapies and systemic psychotherapies where patients can work through the schisms that have arisen in their relationships – should thus be increased as a matter of priority; indeed, in New Zealand there has been an urgent need for increased funding and service reform since well before the pandemic.^61^ Healthcare providers could encourage volunteering^62 63^ and other forms of social prescribing.^64 65^ Public health messaging should highlight the value of repairing interpersonal tensions that arose during lockdown, disseminate advice on how to protect relationships from potentially destructive differences of opinion, and openly acknowledge that ‘returning to normal’ following the challenges of both lockdowns and a global pandemic may need to be undertaken consciously and effortfully, rather than occurring automatically.

Future research in New Zealand should focus on perspectives of people that were not fully captured within our dataset (for example, our study design precluded us from accessing the experiences of under-18s, those too economically disadvantaged to have internet access, and non-English speakers), and the extent to which vaccine rollout can support social recovery. More research is also needed on Māori and Pacific experiences, given the relatively small numbers participating in the survey, the long-standing structural inequities and, relatedly, health disparities affecting these groups, and their disproportionate vulnerability to COVID-19.^43 66-69^ Investigating whether comparable patterns are observed in settings that have achieved elimination in different ways – such as Taiwan, which used a sophisticated contact tracing system rather than lockdowns^70^ – would usefully inform decisions over what *kind* of elimination strategy governments should aspire to in their pandemic planning.

## Conclusion

Where possible – and recognising that the capacity to do so may be limited by the resources available to any given nation-state, as well as by the pathogen’s epidemiology – nations should consider elimination strategies of pandemic response on the grounds that they can enable social recovery, as well as guarding against excess mortality and limiting economic damage. Nevertheless, pandemic planning must anticipate the challenges that certain members of the public might encounter in transitioning back to a satisfying life, especially if elimination requires a period of lockdown or sheltering in place. It is imperative to recognise that pandemic control measures can strain social relationships in various ways, and to furnish populations with resources that can help to resolve such difficulties. Clear guidance on how to socialise safely, and the importance of doing so, is also crucial to ensure concerned members of the public can safeguard their access to social support and thereby protect their own, and others’, physical and mental health.

## Summary Box

### What is already known on this topic

- Elimination strategies have attracted widespread support for their capacity to allow ‘normal’ social and community life to resume.
- There is a paucity of data on how people have been experiencing life in settings where SARS-CoV-2 has been eliminated.

### What this study adds

- Our study shows that while the elimination of SARS-CoV-2 has allowed many people to return to a ‘normal’ social life, such a return is far from inevitable.
- Older people and people with underlying health conditions are especially likely to report becoming less social since the pandemic began, despite the elimination of SARS-CoV-2.
- Elimination strategies should include measures specifically designed to encourage social re-connection.

## Supporting information

Supplemental Annex: Schedule of Survey Questions

SRQR Checklist

## Data Availability

Data underlying the results reported in this manuscript will be available, after deidentification, from the corresponding author upon reasonable request, beginning three months and ending five years following article publication.

## Ethics statements

### Patient Consent for Publication

Not required

### Ethics approval

Ethical approval was provided by the Research Ethics Committee at the London School of Economics and Political Science (ref 11.08c). All respondents were provided with study information before beginning the survey and asked to indicate their consent.

## Data availability statement

Data underlying the results reported in this article will be available, after deidentification, from the corresponding author upon reasonable request, beginning three months and ending five years following article publication.

## Footnotes

### Contributors

NJL, NSA, SGD, AD, EF, EH, NMA, RS, and LT conceived and designed the study. NJL and SGD conducted the coding. All authors contributed to the interpretation of data. NJL conducted the statistical analysis and wrote the first draft. All authors revised the manuscript for intellectual content. Though not involved in designing this study, Pounamu Jade Aikman and Michael Roguski provided valuable input on how to present issues pertaining to ethnicity in the final draft. The corresponding author attests that all listed authors meet authorship criteria and that no others meeting the criteria have been omitted. NJL is guarantor.

### Transparency statement

The lead author (NJL) affirms that the manuscript is an honest, accurate, and transparent account of the study being reported; that no important aspects of the study have been omitted; and that any discrepancies from the study as planned (and, if relevant, registered) have been explained.

### Dissemination to participants and related patient and public communities

There are plans to disseminate the results of the research to the general public, primarily through media outreach (e.g., press releases by the research institutions of the contributing authors, and plain language publications in popular and social media). Findings will also be communicated to participants who provided us with a contact email address.

