## Supplemental Annex: Schedule of Survey Questions for "Social relationships and activities following elimination of SARS-CoV-2: a qualitative cross-sectional study"

### Supplementary Annex: Schedule of Survey Questions

---

Start of Block: Default Question Block

Q1.1

**COVID-19 in 2021: Perspectives from New Zealand**

We are conducting a research project about what it's like to live through the coronavirus pandemic, and would love to hear about your recent experiences of life in New Zealand.

This study involves answering a short survey. It usually takes no more than 20 minutes.

The information you provide will be analysed by a small team of academic researchers based at the London School of Economics, Auckland University of Technology, The University of Auckland, The University of Waikato, and Victoria University of Wellington.

By taking part, you agree you have read and understood the following information about the study.

**What is this project about, and do I have to take part?**

This survey is part of a larger study investigating the effects of the COVID-19 pandemic upon people living in New Zealand and elsewhere. The findings will be used to help develop ways to support people during this outbreak and future ones. Participation is open to people over the age of 18 living in New Zealand and is entirely voluntary.

**What will taking part involve?**

You will complete one online survey. You will be asked questions about yourself, your experiences of the pandemic so far, and your feelings about possible future scenarios.
At the end of the survey, we will ask you whether you might be willing to take part in follow-up research in future. If this is something you can help with, we will ask for an email address.

**Will my taking part and my data be kept confidential? Will it be anonymised?**

Unless you provide us with an email address for follow-up research, the data you provide will be wholly anonymous, and your participation will remain confidential even to us. If you do provide us with an email address, we will store this securely and delete it as soon as the project finished. We will not pass it to any third parties. Nobody will be able to identify you from the anonymised data we analyse or from any publications based on this survey.

**How can I withdraw from the study?**

You may stop answering the survey at any time. If you have already submitted your answers, you may withdraw from the study at any time within fourteen calendar days by contacting the Principal Investigator, Dr Nick Long. However, it will not be possible to remove your information from existing data sets once those data sets have been anonymised, or if you did not supply us with a contact email address, because even we would be unable to identify your data.

**Who has reviewed this study?**
This study has undergone ethics review in accordance with the LSE Research Ethics Policy and Procedure.

**Data Protection Privacy Notice**
The LSE Research Privacy Policy can be found at: https://info.lse.ac.uk/staff/divisions/Secretarys- Division/Assets/Documents/Information-Records-Management/Privacy-Notice-for-Research-v1.1.pdf The legal basis used to process the personal data you provide will be Public Task. The legal basis used to process special category personal data will be for scientific and historical research or statistical purposes.
To request a copy of the data held about you please contact:

**What if I have a question or complaint?**
If you have any questions regarding this study please contact the Principal Investigator, Dr Nick Long, on. If you have any concerns or complaints regarding the conduct of this research, please contact the LSE Research Governance Manager via

---

Q1.2 **Consent Statement**
Please click on this statement to proceed

☐ I confirm that I am at least 18 years old, that I live in New Zealand, that I have read the information about the survey, and that I voluntarily agree to take part in this study. (1)

---

Page Break

Q1.3
**We'd like to start with a few questions about you. These will allow us to see how representative our sample is. All questions are optional, so feel free to skip any you'd rather not answer.**

Q1.4 How old are you?

▼ 18 (1) ... 91 or older (74)

Q1.5 What is your gender?

- ○ Man (1)
- ○ Woman (2)
- ○ Non-binary (3)
- ○ Prefer not to say (4)

Q1.6 Which of the following best describes your sexual orientation?

- ○ Heterosexual (straight) (1)
- ○ Homosexual (gay / lesbian) (2)
- ○ Bisexual (3)
- ○ Asexual (4)
- ○ Other (please specify) (5) ________________________________________________
- ○ Prefer not to say (6)

Q1.7 Which ethnicity / ethnicities do you identify with? Select all that apply.

- ☐ Māori (1)
- ☐ Pacific (2)
- ☐ European New Zealand/Pākeha (3)
- ☐ Asian (4)
- ☐ Middle Eastern (5)
- ☐ Latin American (6)
- ☐ African (7)
- ☐ Other (please specify) (8) ________________________________________________
- ☐ Prefer not to say (9)

Q1.8 Which of the following best describes your religious identity?

- Atheist (1)
- Agnostic (9)
- Christian (2)
- Muslim (3)
- Jewish (4)
- Buddhist (5)
- Hindu (6)
- Sikh (7)
- Not religious but spiritual (10)
- Other (please specify) (8) ________________________________________________
- Prefer not to say (11)

Q1.9 What is the highest level of education you have completed?

- No qualifications (1)
- Completed high school (2)
- Undergraduate degree or professional qualification (5)
- Postgraduate degree (6)
- Prefer not to say (7)

Q1.10 What is your current employment status? Select all that apply.

- ◯ Employed (full-time) (1)
- ◯ Employed (part-time) (12)
- ◯ Self-employed (full-time) (2)
- ◯ Self-employed (part-time) (13)
- ◯ Unemployed (3)
- ◯ Looking after the family / home as a full-time job (5)
- ◯ Unpaid carer (6)
- ◯ In education (7)
- ◯ Long-term sick or disabled (8)
- ◯ Retired (9)
- ◯ Other (please specify) (10) ________________________________________________
- ◯ Prefer not to say (11)

---

Page Break

Q1.11 How many people are living in your household at present?

- 1 - it's just me!  (1)
- 2  (2)
- 3  (3)
- 4  (4)
- 5  (5)
- 6 or more  (6)

---

Q1.12 Are the people living in your household at the moment the people you usually live with?

- Yes  (1)
- No, some people have joined because of the lockdown  (2)
- No, some people are absent because of the lockdown  (3)
- No, for reasons unrelated to the lockdown  (4)
- Prefer not to say  (5)

---

Q1.13 Does your bubble currently include anyone living in another household?

- Yes (please give details)  (1) ________________________________________________
- No  (2)
- Prefer not to say  (3)

---

*Display This Question:*

*If What is your current employment status? Select all that apply. = Employed (full-time)*

*Or What is your current employment status? Select all that apply. = Self-employed (full-time)*

*Or What is your current employment status? Select all that apply. = Employed (part-time)*

*Or What is your current employment status? Select all that apply. = Self-employed (part-time)*

Q1.14 What is your job?

________________________________________________________________

Q1.15 It would help our research to know which part of the country you live in. If you feel comfortable doing so, please share your city, district, or postal code

________________________________________________________________

Page Break

Q1.16 Have you had COVID-19?

○ Yes, confirmed by a test (1)

○ No, not that I am aware (2)

○ Not confirmed, but suspected (3)

○ Prefer not to say (4)

---

Q1.17 Has anyone else in your current bubble had COVID-19?

○ Yes, confirmed by a test (1)

○ No, not that I am aware (2)

○ Not confirmed, but suspected (3)

○ Prefer not to say (4)

---

Q1.18 Do you have any underlying conditions that may affect your vulnerability to COVID-19?

○ Yes (please give details if you feel comfortable doing so) (1)
________________________________________________

○ No (2)

○ Don't know (3)

○ Prefer not to say (4)

---

Q1.19 Does anyone else in your current bubble have any underlying conditions that may affect their vulnerability to COVID-19?

- ○ Yes (please give details if you feel comfortable doing so) (1) ________________________________________________
- ○ No (2)
- ○ Don't know (3)
- ○ Prefer not to say (4)

---

Q1.20 Have you entered or left New Zealand at any time since 19th March 2020?

- ○ Yes (1)
- ○ No (2)
- ○ Prefer not to say (3)

---

Q1.21 Which of the following are currently your main sources of information about how New Zealand is handling COVID-19? Select all that apply.

- ▢ National government website (1)
- ▢ Local council website (2)
- ▢ National news (TV, radio, papers, websites) (3)
- ▢ Local news (TV, radio, papers, websites) (4)
- ▢ Social media (5)
- ▢ Friends and family (6)
- ▢ Employer (7)
- ▢ Community leader (8)
- ▢ Other (please specify) (9) ________________________________________________
- ▢ I have not accessed any such information in recent months (10)

End of Block: Default Question Block

Start of Block: Attitudes

Q2.1
**We'd now like to get an overview of your experiences of and attitudes towards different aspects of the pandemic.**

---

Q2.2
Please indicate how strongly you agree or disagree with each of the following statements:

---

Q2.3 Living in Level 4 lockdown was an unpleasant experience for me.

- ○ Strongly agree (1)
- ○ Somewhat agree (2)
- ○ Neither agree nor disagree (3)
- ○ Somewhat disagree (4)
- ○ Strongly disagree (5)
- ○ Not applicable / don't know / prefer not to say (6)

---

Q2.4 Catching COVID-19 would pose a serious risk to my health.

- ◯ Strongly agree (1)
- ◯ Somewhat agree (2)
- ◯ Neither agree nor disagree (3)
- ◯ Somewhat disagree (4)
- ◯ Strongly disagree (5)
- ◯ Don't know / prefer not to say (6)

---

Q2.5 Over the past six months, I have often worried that COVID-19 is circulating in my community undetected.

- ◯ Strongly agree (1)
- ◯ Somewhat agree (2)
- ◯ Neither agree nor disagree (3)
- ◯ Somewhat disagree (4)
- ◯ Strongly disagree (5)
- ◯ Don't know / prefer not to say (6)

---

Q2.6 I feel confident that vaccinations against COVID-19 are safe.

- ○ Strongly agree (1)
- ○ Somewhat agree (2)
- ○ Neither agree nor disagree (3)
- ○ Somewhat disagree (4)
- ○ Strongly disagree (5)
- ○ Don't know / prefer not to say (6)

Page Break

Q2.7 My household finances have been badly affected by the COVID-19 pandemic.

- ○ Strongly agree (1)
- ○ Somewhat agree (2)
- ○ Neither agree nor disagree (3)
- ○ Somewhat disagree (4)
- ○ Strongly disagree (5)
- ○ Don't know / prefer not to say (6)

---

Q2.8 My life has been badly affected by New Zealand's border restrictions.

- ○ Strongly agree (1)
- ○ Somewhat agree (2)
- ○ Neither agree nor disagree (3)
- ○ Somewhat disagree (4)
- ○ Strongly disagree (5)
- ○ Don't know / prefer not to say (6)

---

Q2.9 Overall, New Zealand has handled COVID-19 better than most other countries.

- Strongly agree (1)
- Somewhat agree (2)
- Neither agree nor disagree (3)
- Somewhat disagree (4)
- Strongly disagree (5)
- Don't know / prefer not to say (6)

End of Block: Attitudes

Start of Block: Snap lockdown

Q3.1
**The whole of New Zealand entered Level 4 lockdown at 23:59 on 17th August 2021.**

---

Q3.2
Do you support or oppose the re-introduction of a Level 4 lockdown?

- ○ Strongly support (1)
- ○ Somewhat support (2)
- ○ Neither support nor oppose (3)
- ○ Somewhat oppose (4)
- ○ Strongly oppose (5)
- ○ Prefer not to say (6)

---

Q3.3 How well prepared were you for this Level 4 lockdown?

- ○ Extremely well prepared (1)
- ○ Somewhat well prepared (2)
- ○ Neither well prepared nor unprepared (3)
- ○ Somewhat unprepared (4)
- ○ Extremely unprepared (5)
- ○ Prefer not to say (6)

---

Q3.4
Before this lockdown was announced, how much had you discussed who would be in your bubble if New Zealand returned to Level 4?

- ◯ A great deal (1)
- ◯ A little (2)
- ◯ Not very much (3)
- ◯ Not at all (4)
- ◯ Prefer not to say (6)

Q3.5 Please explain your answers

________________________________________________________________

________________________________________________________________

________________________________________________________________

________________________________________________________________

________________________________________________________________

Page Break

Q3.6
**Imagine that this lockdown ended up lasting several months...**

Q3.7 What, if anything, would you do differently during this lockdown compared to the last Level 4 lockdown?

________________________________________________________________

________________________________________________________________

________________________________________________________________

________________________________________________________________

________________________________________________________________

Q3.8 Are there any additional forms of support you would hope to receive from the government, compared to the last Level 4 lockdown?

________________________________________________________________

________________________________________________________________

________________________________________________________________

________________________________________________________________

________________________________________________________________

**End of Block: Snap lockdown**

Start of Block: Travel and MIQ

*Display This Block:*

*If Have you entered or left New Zealand at any time since 19th March 2020? = Yes*

Q4.1
**Earlier, you indicated that you had travelled into or out of New Zealand during the pandemic.**

Where did you travel to / from, and for what reasons?

________________________________________________________________

________________________________________________________________

________________________________________________________________

________________________________________________________________

________________________________________________________________

Q4.2 Did any of your travel require you to stay in Managed Isolation and Quarantine (MIQ) facilities?

- ◯ Yes (1)
- ◯ No (2)
- ◯ Prefer not to say (3)

Q4.3 How did you find the experience of travelling during the pandemic?
We are interested in hearing about positive and negative aspects of your travel experience (including your stay in MIQ facilities, where applicable).

________________________________________________________________

________________________________________________________________

Q4.4 What changes could be made to better support people wishing to travel out of, or into, New Zealand during the pandemic?

End of Block: Travel and MIQ

Start of Block: Borders

Q5.1
**We would now like to ask some more about border controls.**

*Display This Question:*

*If My life has been badly affected by New Zealand's border restrictions. = Strongly agree*

*Or My life has been badly affected by New Zealand's border restrictions. = Somewhat agree*

Q5.2 Earlier, you indicated that you had been badly affected by New Zealand's border restrictions. Could you tell us more about how the border restrictions have affected you?

________________________________________________________________

________________________________________________________________

________________________________________________________________

________________________________________________________________

________________________________________________________________

Q5.3 How, if at all, have other people that you know been affected by New Zealand's border restrictions?

________________________________________________________________

________________________________________________________________

________________________________________________________________

________________________________________________________________

________________________________________________________________

Q5.4 What more could the New Zealand government do to support people whose lives are being badly affected by New Zealand's border restrictions?

________________________________________________________________

________________________________________________________________

Page Break

Q5.5 On a scale of 0 to 5, how detailed is your knowledge of New Zealand's current strategy for reopening its borders?

- ○ 0 - I don't know anything about this (1)
- ○ 1 - I know very little about this (2)
- ○ 2 - I know a little about this (3)
- ○ 3 - I know a reasonable amount about this (4)
- ○ 4 - I know a lot about this (5)
- ○ 5 - I know a very great deal about this (6)
- ○ Prefer not to say (7)

---

Page Break

*Display This Question:*

*If On a scale of 0 to 5, how detailed is your knowledge of New Zealand's current strategy for reopen... = 3 - I know a reasonable amount about this*

*Or On a scale of 0 to 5, how detailed is your knowledge of New Zealand's current strategy for reopen... = 4 - I know a lot about this*

*Or On a scale of 0 to 5, how detailed is your knowledge of New Zealand's current strategy for reopen... = 5 - I know a very great deal about this*

Q5.6 How satisfied are you with the government's current plans for reopening New Zealand's borders?

- ○ Very satisfied (1)
- ○ Somewhat satisfied (2)
- ○ Neither satisfied nor dissatisfied (3)
- ○ Somewhat dissatisfied (4)
- ○ Very dissatisfied (5)
- ○ Don't know / prefer not to say (6)

---

*Display This Question:*

*If On a scale of 0 to 5, how detailed is your knowledge of New Zealand's current strategy for reopen... = 3 - I know a reasonable amount about this*

*Or On a scale of 0 to 5, how detailed is your knowledge of New Zealand's current strategy for reopen... = 4 - I know a lot about this*

*Or On a scale of 0 to 5, how detailed is your knowledge of New Zealand's current strategy for reopen... = 5 - I know a very great deal about this*

Q5.7 What, if anything, do you think should be done differently?

________________________________________________________________

________________________________________________________________

________________________________________________________________

________________________________________________________________

---

Page Break

Q5.8 **To what extent do you agree with the following statements:**

Q5.9 New Zealand's borders should remain closed until at least the end of 2021.

- Strongly agree (1)
- Somewhat agree (2)
- Neither agree nor disagree (3)
- Somewhat disagree (4)
- Strongly disagree (5)
- Prefer not to say (6)

Q5.10 New Zealand's borders should remain closed until COVID-19 is under control everywhere in the world.

- Strongly agree (1)
- Somewhat agree (2)
- Neither agree nor disagree (3)
- Somewhat disagree (4)
- Strongly disagree (5)
- Prefer not to say (6)

Q5.11 Athletes should be allowed to travel in and out of New Zealand to take part in international sporting events.

- Strongly agree (1)
- Somewhat agree (2)
- Neither agree nor disagree (3)
- Somewhat disagree (4)
- Strongly disagree (5)
- Prefer not to say (6)

---

Q5.12 The capacity of New Zealand's Managed Isolation and Quarantine (MIQ) facilities should be expanded.

- Strongly agree (1)
- Somewhat agree (2)
- Neither agree nor disagree (3)
- Somewhat disagree (4)
- Strongly disagree (5)
- Prefer not to say (6)

---

Q5.13 When the borders re-open, only fully vaccinated people should be allowed to enter New Zealand.

- ◯ Strongly agree (1)
- ◯ Somewhat agree (2)
- ◯ Neither agree nor disagree (3)
- ◯ Somewhat disagree (4)
- ◯ Strongly disagree (5)
- ◯ Prefer not to say (6)

---

Q5.14 If you would like to, please explain your answers

________________________________________________________________

________________________________________________________________

________________________________________________________________

________________________________________________________________

________________________________________________________________

End of Block: Borders

Start of Block: Before and after

Q6.1

**We would now like to ask about some other ways in which your life may have changed since the start of the COVID-19 pandemic.**

---

Q6.2 Over the past six months, have your **relationships with people who were in your lockdown bubble** been similar or different to how they were before the pandemic?

○ More or less the same (1)

○ A little different (2)

○ Extremely different (3)

---

Q6.3 Please explain your answer

________________________________________________________________

________________________________________________________________

________________________________________________________________

________________________________________________________________

________________________________________________________________

---

Q6.4 Over the past six months, have your **friendships and social life** been similar or different to how they were before the pandemic?

○ More or less the same (1)

○ A little different (2)

○ Extremely different (3)

---

Q6.5 Please explain your answer.

________________________________________________________________

________________________________________________________________

________________________________________________________________

________________________________________________________________

________________________________________________________________

Page Break

Q6.6 Over the past six months, has the current **atmosphere in your local community / neighbourhood** been similar or different to how it was before the pandemic?

- More or less the same (1)
- A little different (2)
- Extremely different (3)

---

Q6.7 Please explain your answer.

________________________________________________________________

________________________________________________________________

________________________________________________________________

________________________________________________________________

________________________________________________________________

---

*Display This Question:*

*If What is your current employment status? Select all that apply. = Employed (full-time)*

*Or What is your current employment status? Select all that apply. = Self-employed (full-time)*

*Or What is your current employment status? Select all that apply. = Employed (part-time)*

*Or What is your current employment status? Select all that apply. = Self-employed (part-time)*

Q6.8 Over the past six months, has your **working life** been similar or different to how it was before the pandemic?

- More or less the same (1)
- A little different (2)
- Extremely different (3)

---

*Display This Question:*

*If What is your current employment status? Select all that apply. = Employed (full-time)*

*Or What is your current employment status? Select all that apply. = Employed (part-time)*

*Or What is your current employment status? Select all that apply. = Self-employed (full-time)*

*Or What is your current employment status? Select all that apply. = Self-employed (part-time)*

Q6.9 Please explain your answer.

________________________________________________________________

________________________________________________________________

________________________________________________________________

________________________________________________________________

________________________________________________________________

Q6.10 Over the past six months, has your **outlook on life** been similar or different to how it was before the pandemic?

- ◯ More or less the same (1)
- ◯ A little different (2)
- ◯ Extremely different (3)

Q6.11 Please explain your answer.

________________________________________________________________

________________________________________________________________

________________________________________________________________

________________________________________________________________

________________________________________________________________

**End of Block: Before and after**

Start of Block: Vaccination

Q7.1
**We would now like to ask you some questions regarding vaccinations against COVID-19.**

---

Q7.2 What are your personal plans regarding vaccination?

- ○ I have already had at least one dose of vaccine (1)
- ○ I plan to get vaccinated as soon as I am able (2)
- ○ I am not sure whether to get vaccinated (3)
- ○ I am not planning to get vaccinated (4)
- ○ Prefer not to say (5)

---

Q7.3 In your opinion, how important is it that people in New Zealand be offered a choice as to which COVID-19 vaccine they receive?

- ○ Extremely important (1)
- ○ Very important (2)
- ○ Moderately important (3)
- ○ Not very important (4)
- ○ Not at all important (5)
- ○ Prefer not to say (6)

---

Q7.4 In your opinion, how important is it that New Zealand secures enough supplies of vaccines for every citizen to be offered multiple booster doses during 2022?

- ◯ Extremely important (1)
- ◯ Very important (2)
- ◯ Moderately important (3)
- ◯ Not very important (4)
- ◯ Not at all important (5)
- ◯ Prefer not to say (6)

---

Q7.5 If you would like to, please explain your answers

________________________________

________________________________

________________________________

________________________________

________________________________

---

Page Break

Q7.6 Do you agree with the following statements:

Q7.7 Vaccinations against COVID-19 should be made compulsory in New Zealand.

- Yes, for everyone who is eligible (unless they are medically exempt) (1)
- Only for certain groups of people (2)
- No (3)
- Don't know (4)
- Prefer not to say (5)

Q7.8 Employers in New Zealand should be allowed to insist that their employees be vaccinated against COVID-19.

- Yes (1)
- Only for certain sectors (2)
- No (3)
- Don't know (4)
- Prefer not to say (5)

Q7.9 Businesses in New Zealand should be allowed to turn away unvaccinated customers and clients.

- ○ Yes (1)
- ○ Only for certain sectors (2)
- ○ No (3)
- ○ Don't know (4)
- ○ Prefer not to say (5)

---

Q7.10 If you would like to, please explain your answers.

________________________________________________________________

________________________________________________________________

________________________________________________________________

________________________________________________________________

________________________________________________________________

**End of Block: Vaccination**

Start of Block: Future scenarios

Q8.1
**Finally, we would like to ask you about a possible future scenario.**

First, imagine that everyone in New Zealand who wishes to be fully vaccinated already has been. COVID-19 cases are then detected in the community.

Q8.2 What changes (if any) would you make to your everyday life in such a scenario, and why?

________________________________________________________________

________________________________________________________________

________________________________________________________________

________________________________________________________________

________________________________________________________________

Q8.3 What action should the government take in such a scenario, and why?

________________________________________________________________

________________________________________________________________

________________________________________________________________

________________________________________________________________

________________________________________________________________

End of Block: Future scenarios

Start of Block: Other countries

Q9.1 Earlier, you indicated that you did not feel New Zealand had the pandemic better than most other countries.

Which countries have handled the pandemic better than (or as well as) New Zealand, and why?

________________________________________________________________

________________________________________________________________

________________________________________________________________

________________________________________________________________

________________________________________________________________

**End of Block: Other countries**

---

**Start of Block: End**

Q10.1 You're almost at the end of the survey. Before you finish, are there any other insights you would like to share with us about your experiences of living through the pandemic in 2021 - or your hopes and fears for the future?

________________________________________________________________

________________________________________________________________

________________________________________________________________

________________________________________________________________

________________________________________________________________

*Display This Question:*

*If Contact List ExternalDataReference Is Empty*

Q10.2 Thank you so much for taking the time to fill in this survey. Your answers will really help us understand how people in New Zealand are experiencing this current phase of the pandemic.

Would you be happy for us to contact you about future opportunities to take part in research on how your life has been affected by COVID-19?

- ◯ Yes (1)
- ◯ No (2)

---

*Display This Question:*

*If Thank you so much for taking the time to fill in this survey. Your answers will really help us un... = Yes*

Q10.3 Fantastic - thank you!

Please let us know the best email address for us to contact you

________________________________________________________________

**End of Block: End**
